# A Novel COVID-19 Early Warning Tool: Moore Swab Method for Wastewater Surveillance at an Institutional Level

**DOI:** 10.1101/2020.12.01.20238006

**Authors:** Pengbo Liu, Makoto Ibaraki, Jamie VanTassell, Kelly Geith, Matthew Cavallo, Rebecca Kann, Christine Moe

## Abstract

SARS-CoV-2 is a respiratory virus but it is also detected in a significant proportion of fecal samples of COVID-19 cases. Recent studies have shown that wastewater surveillance can be a low-cost tool for management of COVID-19 pandemic and tracking COVID-19 outbreaks in communities but most studies have been focusing on sampling from wastewater treatment plants. Institutional level of wastewater surveillance may serve well for early warning purposes since cases can be tracked and immediate action can be executed in the event of positive signal. In this study, a novel Moore swab method was developed and used for wastewater surveillance of COVID-19 at institutional level. Among the 219 swab samples tested, 28 (12.8%) swabs collected from the three campuses and two buildings were positive for SARS-CoV-2. Further individual clinical diagnosis validated the wastewater results and indicated that this method was sensitive enough to detect 1-2 cases in a building. In addition, comparison between grab and Moore swab methods from the hospital sewage line indicated that Moore swab method was more sensitive than the grab sampling method. These results suggest that the Moore swab is a sensitive, practical, and easy to use early warning tool for COVID-19 surveillance especially in low-resource settings and at an early stage of infection in communities.

## INTRODUCTION

Since a novel coronavirus (caused COVID-19) was identified in Wuhan, China in December 2019,^1^ this virus has quickly spread to many countries. The COVID-19 is primarily transmitted person to person via respiratory droplets; however, recent evidence shows that SARS-CoV-2 is also shed in feces. A number of studies have reported that SARS-CoV-2 was detected in a considerable proportion (25%-80%) of fecal samples from COVID-19 cases in both adults and children.^2-6^ In addition, a high proportion of cases had persistently positive viral tests from rectal swabs even after nasopharyngeal swab results became negative, ^2, 3, 5^ suggesting that the duration of virus shedding in the gastrointestinal tract could be longer than the respiratory tract. SARS-CoV-2 shedding in the gastrointestinal tract enables the detection of SARS-CoV-2 in wastewater, which have been reported by Wu et al.^7^ This study^7^ reported the detection of high titers of SARS-CoV-2 in samples from an urban wastewater treatment plant that were collected in mid- to late-March 2020, but analyses of archived sewage samples from January 2020 were negative. This study suggests that the detection of SARS-CoV-2 can be an early warning signal of COVID-19 epidemics in the community.

Wastewater surveillance has been traditionally performed using two methods: grab sampling and composite sampling. Grab sampling is a simple and convenient method that involves filling a container with wastewater at one point in time; however, interpretation is limited because samples only represent a snapshot at one moment. Composite sampling has been considered as a more representative method due to its ability to collect numerous individual samples at regular intervals and individual samples are combined in proportion to the wastewater flow rate.^8^ Composite samples can be performed manually or by using automated samplers, such as flow-weighted samplers or continuous composite samplers. Automated composite samplers must have built-in refrigeration capacity and require electricity, whereas manual composite samples require samplers to repeatedly return to the same location multiple times over a period of time. Composite sampling is, therefore, more costly and time-consuming, and it may not be feasible under certain environmental conditions (e.g. no electrical hook-ups) or when resources are limited, but composite samples are more appropriate for analysis over grab samples.

Wastewater surveillance for SARS-CoV-2 can be performed in the downstream or upstream of sewage line.^9^ Sampling untreated influent or effluent from wastewater treatment plants is a useful method to evaluate community-level trends of COVID-19 infection but the catchment area contributing sewage to this point will be large since wastewater treatment plants in urban areas may serve tens of thousands of people. Alternatively, wastewater surveillance from the upstream of these plants can be used to detect SARS-CoV-2 in a smaller catchment. For example, collecting wastewater samples from manholes at an institution-level provides researchers and decision-makers better understanding SARS-CoV-2 presence in wastewater on the premises.

The “Moore swab” method is an environmental surveillance method that has been used for decades by public health practitioners around the world to detect and isolate enteric pathogens from water. Consisting of a strip of gauze tied with string suspended in flowing water, this sampling method acts as a filter that allows collection of microorganisms over an extended period of time. The Moore swab was first proposed by Brendan Moore in 1948 to trace *Salmonella Paratyphi B* from effluent sewage water in North Devon, England to determine the sources of infection responsible for sporadic outbreaks of paratyphoid fever.^10^ Since the first application to detect *typhoidal Salmonella* in sewage water, the Moore swab method has been utilized throughout the world to detect several fecal borne pathogens such as coxsackieviruses,^11^ poliovirus (26-29),^12-15^ human norovirus,^16^ *Escherichia coli* O157:H7 ^16, 17^ *Vibrio cholerae* O1.^18-20^ Over the years, the technique once used for tracing the chronic carriers of *Salmonella Paratyphi B* has expanded to environmental surveillance, investigation of ongoing outbreaks, and bacterial enumeration in surface water.^10^

The objectives of this study were to develop a practical, low-cost, and convenient Moore swab method for composite wastewater sample collection and provide early warning of COVID-19 outbreaks at the institutional level. A positive PCR signal from a Moore swab placed in wastewater flowing by a building can be used to help inform clinical surveillance and initiate diagnostic testing of residents in buildings. For these objectives, we were interested in the presence or absence of SARS-CoV-2 in sewage line instead of quantitative results.

## MATERIALS & METHODS

The Moore swab method includes five steps: swab sample collection, laboratory processing, polyethylene glycol (PEG) precipitation, RNA extraction and quantitative Real-time RT-PCR (RT-qPCR) detection of SARS-CoV-2 viruses.

### Moore Swab Sample Collection and Processing

The Moore swabs were made by cutting pieces of cotton gauze approximately 120 cm long by 15 cm wide and firmly tying the center with fishing line. For sample collection, a Moore swab was placed in the outflow stream of the wastewater from buildings to be tested, secured by tying a fishing line to a hook at the top of the manhole. After leaving it in place for 24-72 hours, the swab was collected from the wastewater, stored in a Ziploc bag, and transported to the lab on ice. Samples were placed in a cooler during transportation to prevent possible overheating. For swab processing, the swab was squeezed in a beaker to get all the trapped liquid out, which was then poured into a flask. The swab was then submerged in 100 mL elution buffer ^21^ while gently kneading the swab for 2 minutes. The liquid from the swab was merged with the initial squeezed sample inside the flask. This step was repeated two more times until the total flask volume was approximately 300 mL. The swab was then disposed and the liquid in the flask was poured back into the initial beaker. A piece of double-layered gauze was placed over the opening of the flask, and the liquid in the beaker was poured through to filter out large solid materials until a final sample volume of 250 mL was collected. Finally, the liquid was centrifuged at 6,000 rpm for 15 minutes at 4°C to remove additional solids, and the supernatant was saved for further processing.

### Grab sample collection and processing

A grab sample was collected from the outflow stream of the sewage from buildings to be tested. 1 L of sewage water was collected in a sterile, autoclavable, polypropylene bottle. Collection of grab samples was only possible for sewage outflow with adequate flow. Grab samples were transported to the lab in a cooler with ice to prevent possible overheating. The sample was first pasteurized at 65°C for 1 hour to inactivate pathogens before sample processing. 500 mL of the sample was centrifuged at 6,000 rpm for 15 minutes at 4°C to remove solids in the sample that had potential of clogging the membrane filter. The sample was processed by performing membrane filtration with a sample volume conducive to filtration in about 3-5 hours depending on turbidity. The sample was filtered through two 0.45-μm-pore-size, 47-mm-diameter nitrocellulose filters (Millipore Sigma, Burlington MA) in order to maximize filtration amount after pH adjustment to 3.5 and addition of 10^5^ of Bovine Respiratory Syncytial Virus (BRSV) (INFORCE 3, Zoetis, Parsippany, NJ). After filtration, the membrane filter was placed into a microcentrifuge tube and buffer RLT from the RNeasy Mini Kit was added immediately. The sample was subjected to RNA extraction described later in the paper after it was vortexed at maximum speed for 10 minutes.

### PEG Precipitation

All Moore swab samples were processed by adding 12% PEG (Sigma, St. Louis, MO, USA), 0.9 Mole sodium chloride, and 1% bovine serum albumin (Sigma, St. Louis, MO, USA).^21^ To monitor RNA extraction procedure and the PCR inhibition of SARS-CoV-2, 10^5^ of BRSV were added into each sample as an extraction blank control prior to PEG precipitation. A stir bar was added to each flask and the samples were spun overnight in a cold room.

### RNA Extraction

After ∼16 hours of PEG precipitation, samples were centrifuged at 6,000 rpm for 1 hour at 4°C. Supernatant was removed along with the bulk of the pellet to reduce excess waste material and improve extraction yield. Walls of the centrifuge bottle were rinsed and remaining solids were dissolved using 800 μl RLT buffer from the RNeasy Mini Kit (QIAGEN, Hilden, Germany) following the protocol. Finally, 100 μl of RNA was achieved from each sample.

### Quantitative Real-Time RT-PCR Method

SARS-CoV-2 RNA were detected via real-time quantitative reverse-transcription polymerase chain reaction (RT-qPCR) using the N1 and N2 primers developed by CDC.^22^ Inactivated SARS-CoV-2 RNA (ATCC, Manassas, VA) served as a positive control. SARS-CoV-2 RNA titers for samples were estimated based on interpolation with a standard amplification curve for the positive control. BRSV was detected using the primers/probe described by Boxus et al.^23^ A one-step RT-qPCR was successfully developed and validated using 10-fold serial diluted SARS-CoV-2 RNA standard. The Invitrogen SuperScript III One-Step RT-PCR with Platinum Taq kit was used (Invitrogen, Carlsbad CA). Each reaction contained 1 μL of SuperScript III RT/Platinum Taq Mix, 1x Reaction Mix, and 0.04 mM Magnesium Sulfate from the kit. Additionally, each reaction contained 0.2 μM probes and 0.04 μM primers specific to either SARS-CoV-2 or BRSV. 5 μL of RNA sample was loaded in duplicates into a 96-well plate with the reaction mixture and placed into the Bio-Rad CFX PCR thermocycler (Bio-Rad, Hercules CA). The RT-qPCR program consisted of 25°C for 2 minutes, 55°C for 13 minutes, 95°C for 3 minutes, 95°C for 15 seconds, and 58°C for 30 seconds, with a total of 45 cycles. Cycle threshold (Ct) values were recorded for each reaction.

### Moore Swab Seeding Experiment

A Moore swab was placed in a plastic container filled with 2 L distilled water and the end of the fishing line was attached to a shelf located above the container. A stir bar was placed in the container to mimic current observed in the field. The distilled water was spiked with 50 genome equivalent copies (GEC)/mL, 5 GEC/mL, and 0.5 GEC/mL of known amounts of inactivated SARS-CoV-2 (ATCC, Manassas, VA) and BRSV. The swab was submerged in the flowing water for 24 to 72 hours to trap the inactivated SARS-CoV-2 and BRSV as shown in Figure 1. The swabs were subjected to RNA extraction as described in the paper.

**Figure 1:**
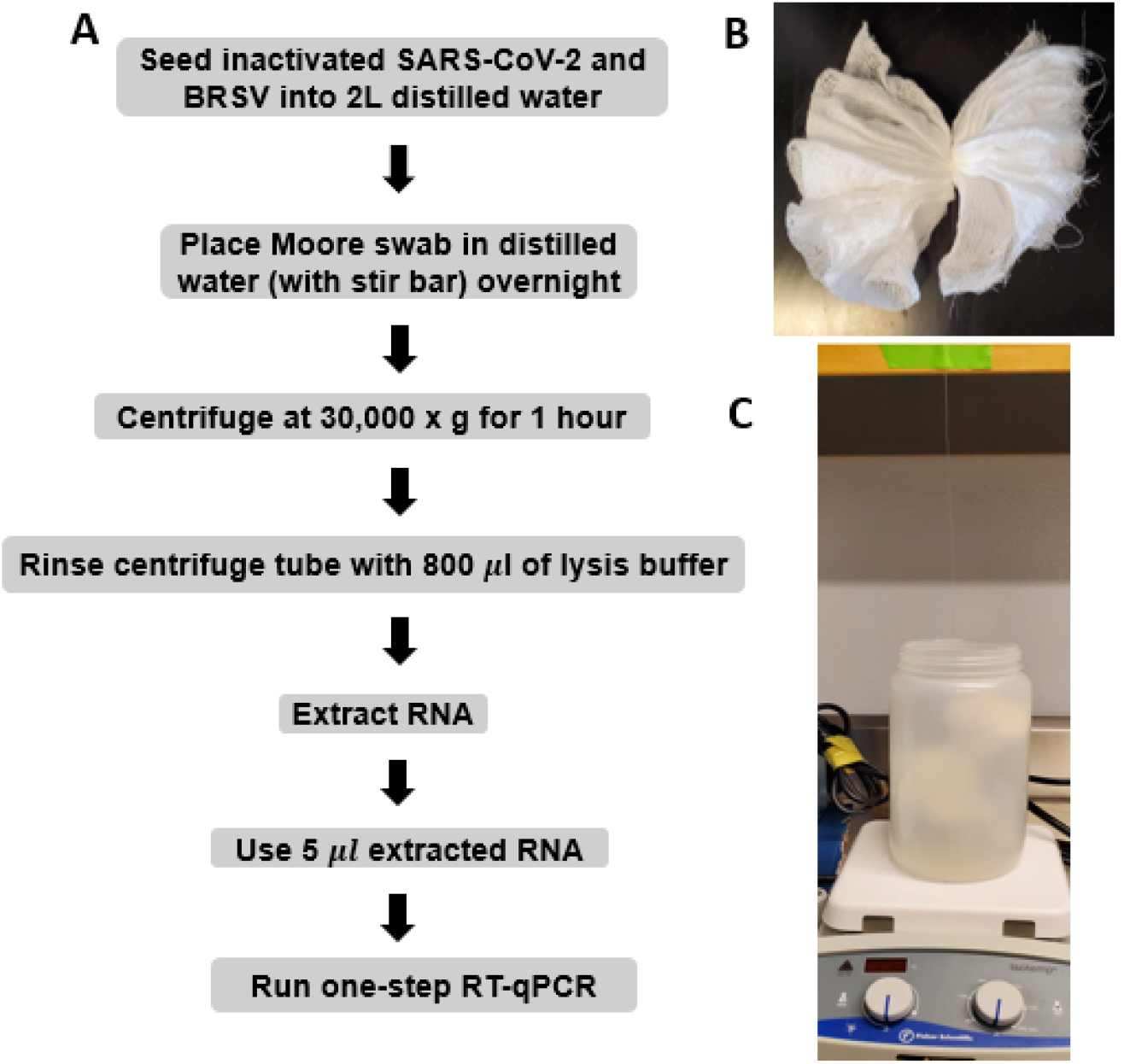
Seeding experiment of distilled water seeded with inactivated SARS-CoV-2 and BRSV. A). Flow diagram showing Moore swab samples processing and testing procedures; B). Moore swab; C). Seeding experiment setup.

## RESULTS

### 1. Moore swab method for recovering seeded SARS-CoV-2 and BRSV in distilled water

When both SARS-CoV-2 and BRSV were seeded at 50 GEC/ml, all three swabs showed positive results using the RT-qPCR methods. When the seeding level was reduced to 5 GEC/ml, SARS-CoV-2 could only be detected in one of the two experiments, but BRSV was detected in both experiments. At the 0.5 GEC/ml seeding level, both SARS-CoV-2 and BRSV could not be detected with the RT-qPCR assays (Table 1), indicating the limit of detection of both SARS-CoV-2 and BRSV using the Moore swab method in 2 liters distilled water was 5 GEC/ml.

**Table 1.** Moore swab method for recovering seeded SARS-CoV-2 and BRSV in distilled water

| **Seeding Level**<br>**(GEC\*/ml)** | **SARS-CoV-2 Positive**<br>**Swabs/no. Swabs** | **BRSV Positive**<br>**Swabs/no. Swabs** |
| --- | --- | --- |
| 50 | 3/3 | 3/3 |
| 5 | 1/2 | 2/2 |
| 0.5 | 0/2 | 0/2 |

\*Genome equivalent copies

### 2. SARS-CoV-2 detection using Moore swabs on Emory campuses

Multiple residence halls at three Emory campuses (Main, Clairmont, and Oxford) were sampled on a weekly basis starting mid-August, 2020 for a total of 219 Moore swab samples. The swab positive rates were 0-16.2% for the three campuses with Main Campus the highest (16.2%) and Oxford the lowest (0%), 46.2% (6/13) swabs from the quarantine building where confirmed COVID-19 cases were isolated, and 91.7% (11/12) swabs from Emory hospital sewage line that weekly Moore swab samples were positive (Table 2).

**Table 2.** SARS-CoV-2 Detection using Moore Swabs on Emory Campus

| **Campus/Building** | **No.**<br>**Manhole** | **Swabs**<br>**Tested** | **Swabs Positive\* (%)** |
| --- | --- | --- | --- |
| Main | 10 | 62 | 10 (16.2) |
| Clairmont | 7 | 76 | 1 (1.3) |
| Oxford | 6 | 56 | 0 (0) |
| Quarantine Bldg | 3 | 13 | 6 (46.2) |
| Emory Hospital | 1 | 12 | 11 (91.7) |
| Total | 27 | 219 | 28 (12.8) |

\* Ct values of positive swabs were between 23.6-39.5

### 3. Detection of SARS-CoV-2 in wastewater using Moore swabs in a quarantine building

Students living in dormitories who tested positive for COVID-19 were moved to a quarantine building near Main Campus. The building has two manhole locations to place Moore swabs that are referred to as North Wing manhole and South Wing manhole. The North Wing manhole contains two pipes, and these are denoted as “Site 1” and “Site 2”. The South Wing only has one pipe that is denoted as “Site 3” (Table 3). For proof of concept purpose, Moore swabs were initially placed in all sites. Swabs from South Wing, where contained confirmed COVID-19 cases, were positive for SARS-CoV-2 in four out of six swab samples (66.7%). Swabs from the North Wing, where there were no known cases, were all negative. On September 29, 2020, a positive swab was detected when there was only one case in the building but this was not true on September 22 and October 6 when there were eight existing cases (Table 3) These results suggest that the Moore swab method is a sensitive and specific technique for SARS-CoV-2 detection in sewage. Beginning on September 22, 2020, the North Wing of the building was removed from the sampling scheme due to our limited resources and supplies.

**Table 3.** Moore Swab SARS-CoV-2 RT-qPCR Detection in Quarantine Building

| Swab Placement Date | Case Number | Site 1 | Site 2 | Site 3 |
| --- | --- | --- | --- | --- |
| 9/8/2020 | 2 | ND* | ND | Positive |
| 9/14/2020 | 5 | ND | ND | Positive |
| 9/22/2020 | 1 | - | - | ND |
| 9/29/2020 | 1 | - | - | Positive |
| 10/8/2020 | 8 | - | - | ND |
| 10/15/2020 | 4 | - | - | Positive |

\*ND: not detected

### 4. SARS-CoV-2 detection in grab and Moore swab samples from Emory hospital sewage

Only grab samples were collected from Emory University Hospital’s sewage line between weeks 1 (July 12, 2020) and 5 (August 9, 2020). Starting from week 6 (August 16, 2020), both grab and Moore swab samples were collected. Grab samples were collected around the same time on Monday mornings while Moore swabs were also collected around the same time on Wednesday mornings. For each week when samples were collected, we also obtained COVID-19 inpatient cases at the Emory University Hospital. 10 out of the 13 grab samples were positive and 7 out of 8 Moore swab samples were positive (Table 4). While there was some inconsistency with the detection of SARS-COV-2 in the wastewater samples throughout the 13 weeks of collection, the sensitivity of Moore swab samples tended to be better than grab samples.

**Table 4.**
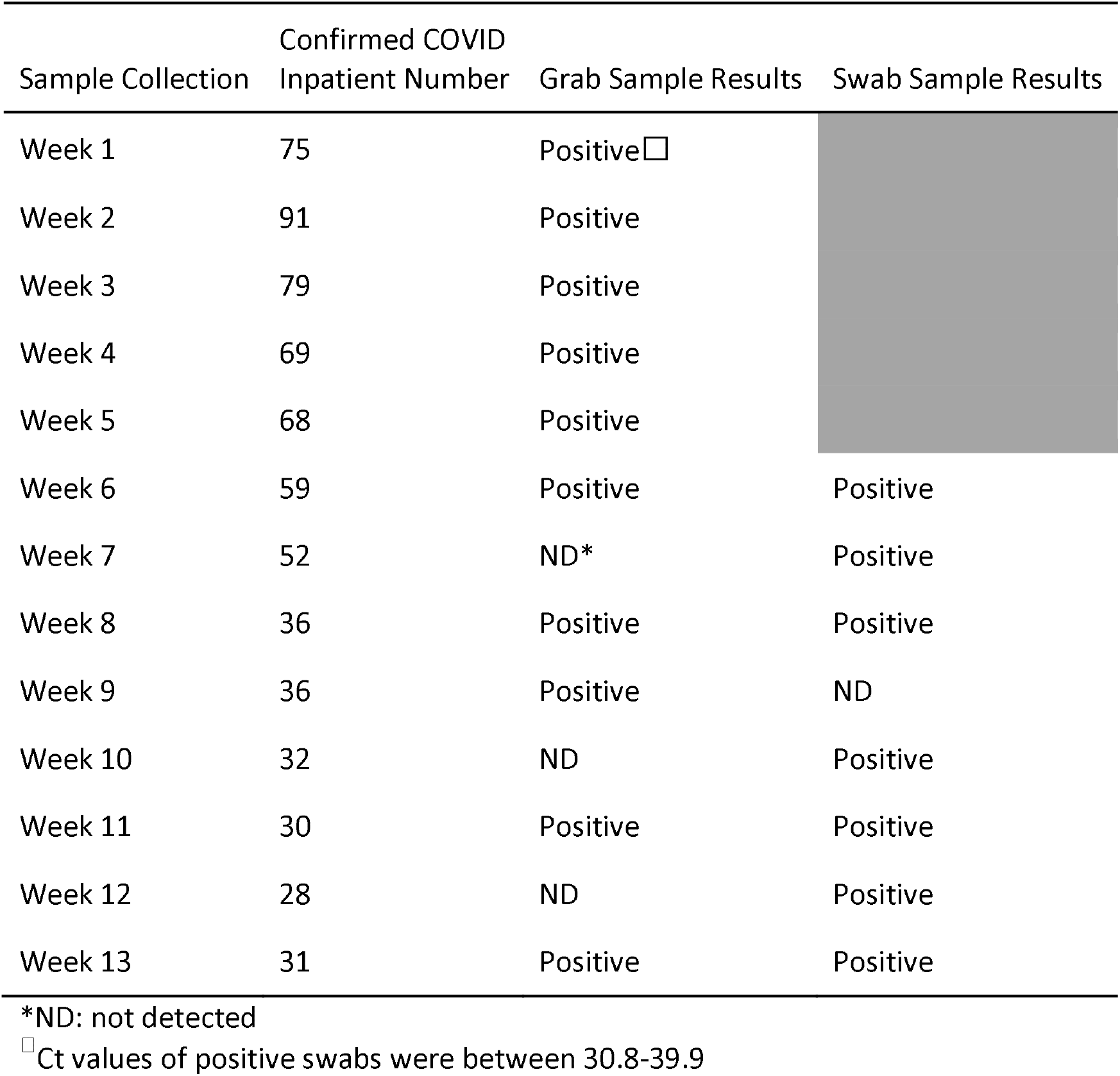
SARS-CoV-2 RT-qPCR Results of Grab and Moore Swab Samples of Emory University Hospital Wastewater and Corresponding COVID-19 Case Numbers, June - October 2020

## DISCUSSION

This study shows proof of concept for the application of the Moore swab sampling as an early warning of presence of COVID-19 infections in the residence halls/buildings on university campuses. In the present study, we were able to develop the Moore swab method for sewage sampling in manholes, laboratory processing of swab samples, viral concentration, RNA extraction, and RT-qPCR detection of SARS-CoV-2 in wastewater. The limit of detection of the Moore swab method, evaluated using inactivated SARS-CoV-2 and BRSV in distilled water, was 5 GEC/mL. Among the 219 swab samples tested, 28 (12.8%) swabs collected from the three campuses and two buildings were positive for SARS-CoV-2. Results from the quarantine building and a student residence hall (data not shown in this study) validated the fact that this method was sensitive enough to detect 1-2 cases in a building. In addition, comparison between grab and Moore swab methods from the hospital sewage line indicated that Moore swab method was more sensitive than the grab sampling method. These results suggest that the Moore swab is a sensitive, practical, and easy to use method and it can be applied at any building level, like schools, nursing homes, prisons, and public/private buildings.

This method is particularly appropriate for COVID-19 prevention and control in low-resource settings and at an early stage of infection in communities since the positive signal from the sewage line could allow deployment of rapid response teams into the building to conduct more intensive diagnostics and isolation of infected individuals to prevent further spread of the virus.

Pathogen detection in wastewater can be a valuable surveillance approach - especially for diseases with pre-symptomatic or asymptomatic transmissions, such as COVID-19. From a surveillance perspective, wastewater surveillance is advantageous because it can monitor the disease epidemic status and provide early warning for potential outbreaks for a relatively large population without having to collect individual clinical specimens, and it captures both symptomatic and asymptomatic infections. In addition, collecting and analyzing wastewater samples is relatively simple, does not require informed consent, is cost-efficient, and can provide rapid results. wastewater surveillance provided an early warning of cholera in Peru in the early 1990’s and has been an integral part of the global polio eradication program for several decades. It has been proven useful in early warning of poliovirus circulation in a range of countries and populations.^24-27^ Studies from several countries ^25-27^ indicated that wastewater surveillance was able to detect wildtype poliovirus in sewage even in the absence of reported acute flaccid paralysis cases. This may also be true for COVID-19 surveillance since recent publications from two countries ^28, 29^ have reported that asymptomatic infections of COVID-19 were very common and estimates of prevalence primarily based on symptomatic individuals testing might be significant underestimations.

Detection of SARS-CoV-2 RNA in untreated wastewater has been reported in a number of countries, such as the Netherlands,^30^ USA,^31^ Australia,^32^ China,^33^ Spain.^34^, and Italy. ^35^All these studies have been focusing on sampling from wastewater treatment plants and positive PCR signal may represent hundreds of thousands of infected cases in the catchment areas. Results from downstream of sewage lines may be used for surveillance purposes or to monitoring the trend of disease prevalence in the catchment population but when the disease prevalence is low or the virus is highly diluted in the sewage system, the method may not be sensitive enough to detect positive signals. On the other hand, when the disease prevalence is high, samples at the downstream tend to have consistent positive signal and cannot locate the infections from the catchment area for targeted population interventions. However, wastewater from the upstream is less diluted, from a smaller catchment area, and could easily interpret results when a population has an increasing number of new infections, which may lead to early warning and early intervention to prevent further transmissions. To date, there have been no published data demonstrating the use of wastewater surveillance to identify SARS-CoV-2 at building level. Institutional level of wastewater surveillance serves well for early warning purposes since immediate action can be executed in the event of any positive signal to prevent possible outbreaks. However, there are several fundamental challenges to sampling a building sewage for SARS-CoV-2 that is transiently present. The challenges include small wastewater flow at the sampling site (e.g, manhole), depth of the manhole, intermittent excretion of viruses, unknown viral peak time, composite sampling, sample time, and frequency. Among these challenges, composite sampling using an automated composite sampler is a good solution to overcome some challenges even though grab sampling is an easy and convenient way for wastewater collection. Composite samplers, however, are not affordable to most researchers, especially in resource limited settings and a huge demand can be anticipated for this expensive equipment after more COVID-19 sewage water surveillances start. We have recently successfully applied Moore swabs for wastewater-based surveillance at the city and neighborhood level for typhoid fever in Kolkata, India and our preliminary data indicate that this method is more sensitive than grab samples (data not published). In this study, we successfully developed and applied the Moore swab method for COVID-19 wastewater surveillance. The Moore swab can act as a composite sampler that can trap microorganisms in water. Given its simplicity and affordability, the Moore swab method is well suited for COVID-19 wastewater surveillance in buildings or communities where COVID-19 prevalence is relatively low and wastewater surveillance is feasible through a sewage network. The identification of positive signal in a building or community may trigger epidemiological investigation to identify those who are infected. Our results showed that the Moore swab method was sensitive enough to identify one or two infected cases in a building.

Although there are advantages with using the Moore swab method, the method also has several limitations. First, this method only provides results on the presence or absence of cases so the quantitative assessment work will be limited. Second, each Moore swab sample requires two trips: placing and retrieving a swab while grab sampling only requires one trip. Third, the Moore swab method has not been standardized in terms of sampling frequency, duration of immersion, and swab processing procedure. Lastly, because this study is a prove of concept, consistent positive results were not achieved in few times when there were confirmed cases in the buildings. This might reflect swab sample processing variability, different RNA extraction methods, laboratory protocol changes, and other unknown factors, which may not represent the true situation.

## Data Availability

All our data are available upon request

## ACKNOWLEDGEMENT

We would like to thank the Emory Maintenance Department, including Raymond Pullins, Matthew Thatcher, and the Oxford Campus staff, for their efforts in identifying and providing access to sewage sampling locations. We would further like to thank the Emory COVID Response Team, including Samuel Shartar, Colleen Kraft, and Morgan Lane, for providing inpatient hospitalization data at Emory hospital, quarantine information, and individual diagnostic results from the student residence halls.

